# Can Blood Gas Enhance Early Warning Systems by Streamlining ICU Transfer Decisions: A Qualitative Systematic Review

**DOI:** 10.1101/2025.10.23.25338637

**Authors:** Praveen Meka, Emma Stiller

**Author notes:** **Corresponding Author:** Emma Stiller.

## Abstract

**Importance:** Delay in transfer to Intensive Care Unit (ICU) is associated with known adverse clinical and economic outcomes. There are several early warning systems (EWS) that help identify patients that could benefit from earlier ICU transfer but are fraught with challenges when used to measure delays. An objective time stamped blood test metric, such as a blood gas analysis (BGA), could be a valuable adjunct in identifying patients and measuring delays in who require intrahospital transfer to the ICU.

**Background:** Delays in transferring critically ill patients to the ICU are linked to increased mortality, organ failure, prolonged recovery, and higher hospital costs. While EWS systems like MEWS, NEWS2, and eCART aim to detect deterioration using vital signs and medical data, they often rely on intermittent and/or subjective inputs. Despite advances, including AI-driven models, most systems still lack accuracy to detect and quantify transfer delays, an important operational metric.

**Objective:** This review explores the clinical and operational impact of ICU transfer delays and evaluates the potential role of BGA as an objective, time-stamped adjunct biomarker for early identification of high-risk patients. We also assess whether BGA could be integrated into EWS tools to enhance predictive accuracy.

**Methods:** We conducted a systematic literature review of studies published between 1994 and 2024 using PubMed, EMBASE, Cochrane, and NIH databases. Inclusion criteria focused on studies that examined ICU transfer delays, BGA parameters (e.g., lactate, pH, base excess), and clinical outcomes in adult or pediatric patients. Studies were excluded if they had small sample sizes (n < 50), lacked outcome data, or were not published in English.

**Results:** The review found that delays in ICU transfer are consistently linked to worse clinical outcomes and higher healthcare costs. While EWS tools have improved early recognition of patient deterioration, they still lack objective, time-stamped markers to measure delays. Approximately one-third of the included studies specifically examined BGA parameters in relation to ICU transfer or outcomes. Elevated lactate levels and abnormal pH values correlated with increased ICU admission, adverse prognosis and mortality risk. Despite this, BGA is not currently integrated into most clinical decision-making tools used for ICU triage.

**Conclusion:** BGA represents a promising, underutilized tool that could fill a critical gap in current ICU triage systems. As a time-stamped, objective measure of physiological instability, BGA could enhance the accuracy, timeliness and measurability of ICU transfer decisions—especially when combined with electronic medical records and modern EWS platforms. Future research should focus on evaluating BGA as a predictive input within next-generation EWS tools, with the goal of reducing ICU transfer delays, improving patient outcomes, and optimizing hospital resource use.

## Introduction

Patients transferred from inpatient wards to ICUs often experience delays that can negatively affect their health and recovery. ICUs offer critical care expertise, higher staff-to-patient ratios, and access to specialized procedures and equipment. However, once a patient is identified as needing ICU care for intrahospital transfer, multiple steps must be taken—including communication among staff, preparation of the ICU room, and readiness of personnel and equipment—before the transfer can occur. These steps can contribute to delays that compromise patient outcomes. Numerous studies support that earlier ICU admission improves recovery and reduces overall hospital length of stay (Churpek et al., 2016). Several studies have explored the causes and effects of ICU transfer delays, as well as tools that may help streamline the process. For instance, risk scoring systems such as the electronic Cardiac Arrest Risk Triage (eCART) score have been shown to aid in early identification of deteriorating patients and support faster clinical decision-making Without such objective tools, physicians may hesitate to escalate care until they are certain ICU-level resources are needed, leading to harmful delays (Gallo et al., 2024).

Barriers to escalation include a lack of standardized protocols, resources, accountability, rapid response behaviors, human factors such as fear, hierarchy, conflict, confidence, and patient factors. These are all individual yet inter-related barriers and facilitators to escalation. In a study, to address these barriers and improve escalation of care according to EWS protocols, delays were reduced into five overarching themes: Governance, Rapid Response Team (RRT) Response, Professional Boundaries, Clinical Experience, and EWS parameters. (Oneil et al., 2021). All these factors combined ultimately created delay in ICU transfer of critically ill which was associated with increased morbidity and mortality (Dunser et al., 2024). ICU transfer delay due to communication flaws and inconsistent transfer guidelines can be partially mitigated with tools such as early warning scores (EWS). EWS tools have demonstrated promise in streamlining ICU transfers. We hypothesize that combining EWS with time stamped BGA may further enhance early identification of patients and measuring delays in those who need ICU care. This objective metric, defined by specific qualifications, would help minimize delays associated with ICU transfers, enabling patients to receive timely care - a factor shown in numerous studies to improve patient recovery outcomes and reduce overall length of stay (Churpek et al, 2016).

A study comparing six EWS tools, found national early warning score (NEWS) the most accurate EWS for predicting the risk of death/ICU admission within 24 hours from ED arrival. To note, the NEWS score was calibrated with few events occurring in patients classified at low risk. Metrics scored included respiration rate, oxygen saturation (SpO2), need for supplemental oxygen, temperature, systolic blood pressure (SBP), heart rate, and level of consciousness (Covino et al. 2023). The evaluation of experience using NEWS/NEWS2 within the national health service (NHS), the first healthcare system globally to adopt a ‘common language’ for illness severity, has shown some success in creating a standardized EWS for acute clinical illness and deterioration. This system is now being replicated in many other areas of the world (William B, 2022). High-risk patients identified by EWS who further experience delays in ICU transfer show higher mortality, and prolonged hospital stays. Accurate, continuous monitoring and documentation of EWS can predict patients who may benefit from early intervention (Gielen et al., 2021).

The objectives of this review are to examine the clinical and operational impact of ICU transfer delays, assess varied available EWS tools, and evaluate BGA as a time-sensitive biomarker and predictive tool to enhance early risk stratification and facilitate ICU transfers.

## Methods

The PRISMA (Preferred Reporting Items for Systematic and Meta-Analyses) statement for reporting was used for this study (Figure 1 & Figure 2). An exhaustive literature search of literature was conducted using the following databases: PubMed, EMBASE, Cochrane, and the National Institutes of Health (NIH) database. These comprehensive databases were selected to ensure a thorough and systematic review of the available literature. The search was limited to publications from 1994 to 2024, covering a 30-year period to capture the most relevant and up-to-date research on the topic.

**Figure 1:**
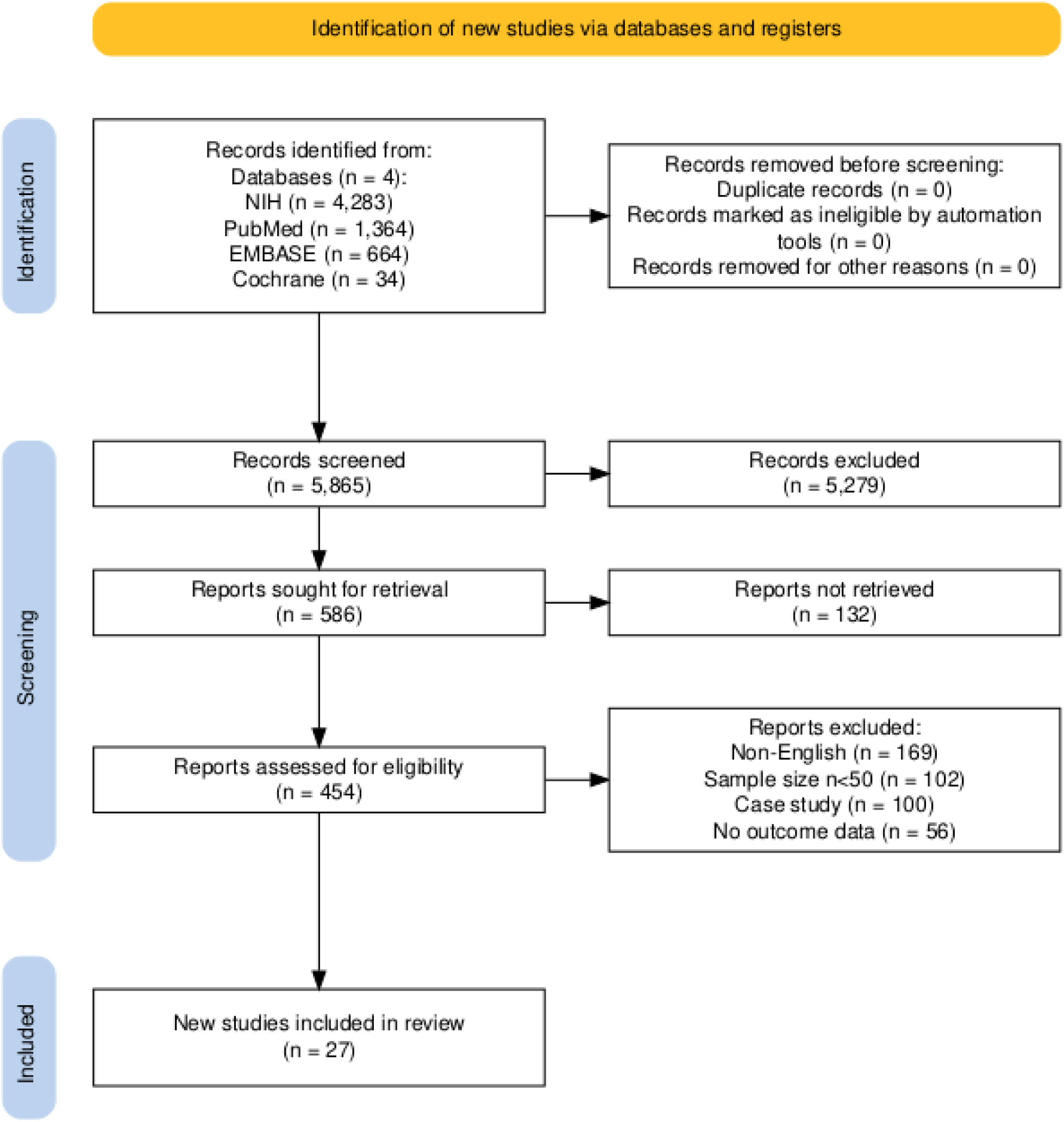
Evaluating clinical outcomes as a result of delay in ICU transfer.

**Figure 2:**
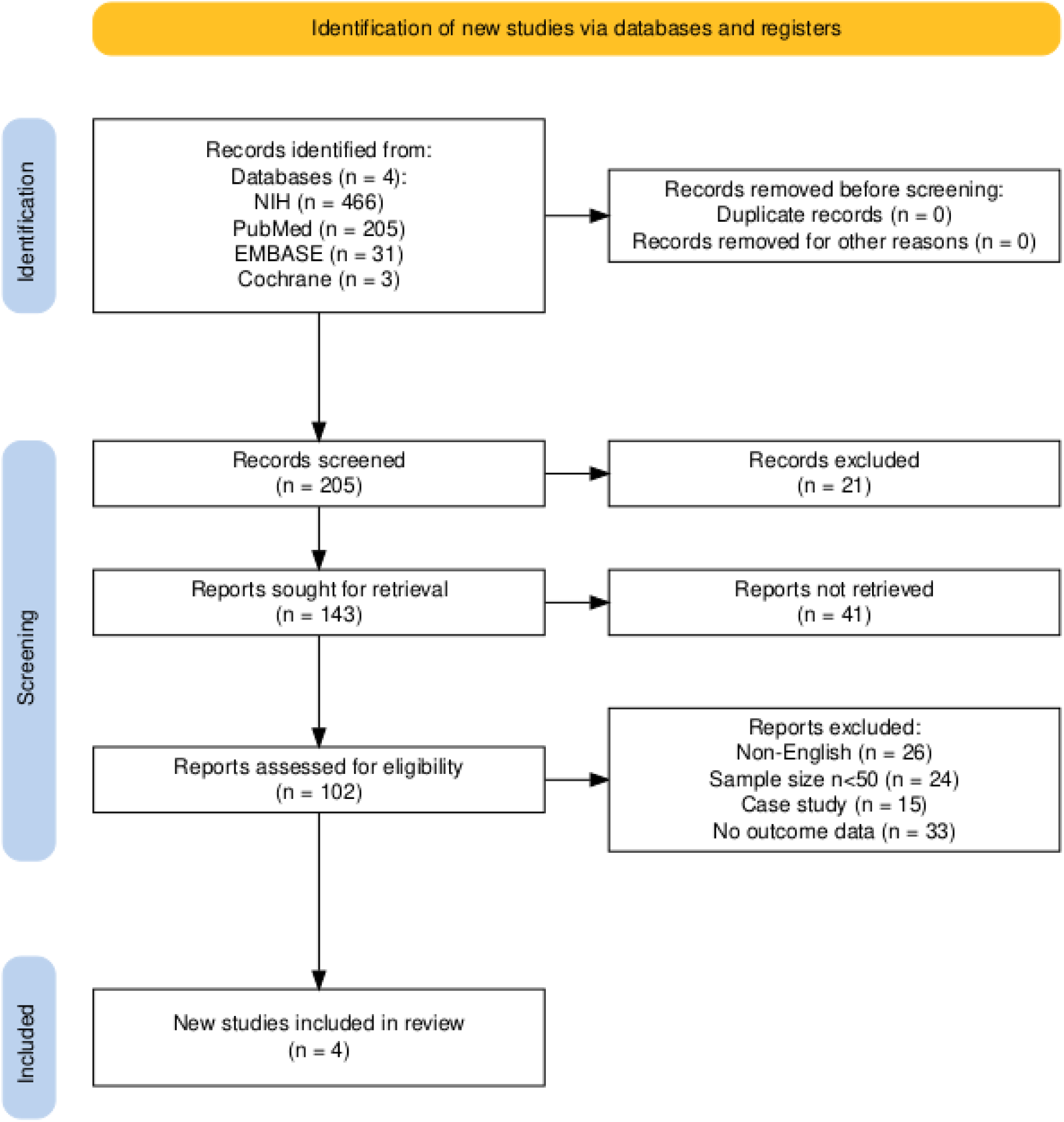
Evaluating economic outcomes as a result of delay in ICU transfer.

### Study Selection

The main author performed keyword search followed by abstract review followed by full text-review of qualified articles. The search keywords used were: “ICU transfer/admission”, “delayed transfer”, “critical care”, “blood gas analysis”, “patient outcomes”, “in-hospital mortality”, “treatment delay”, and “length of stay”. The keywords identified studies that examined the impact of ICU transfer delays and associated clinical outcomes.

The *inclusion criteria* for the qualitative review were studies examining ICU transfer delays, studies reporting blood gas parameters, adult and pediatric populations, English language publications, and primary research articles. The inclusion of both adult and pediatric populations was a deliberate methodological decision, justified by the fact that BGA constitutes a common diagnostic modality indicated by similar needs across all age groups and reflects shared underlying respiratory and metabolic physiology, despite anticipated age-related variations in reference values.

The *exclusion criteria* were for the qualitative review were studies with a sample size of less than fifty (n=126), case reports (n=115), non-English publications (n=195), and studies without outcome data (n=89).

Application of these comprehensive search strategies and inclusion/exclusion criteria allowed the authors to identify and synthesize high-quality evidence on the impact of ICU transfer delays, components of blood gas as predictors, and associated clinical outcomes.

### Data Synthesis

The extracted data was synthesized qualitatively to allow comprehensive understanding of the relationship between ICU transfer delays and patient outcomes. The data was analyzed to identify patterns and trends, including the impact of delayed transfers on in-hospital mortality, treatment delay, and length of stay. Key findings and implications were highlighted relevant to clinical practice. Biases such as selectivity bias or confounder bias were corrected on an ongoing basis when identified.

### Outcomes Studied

The primary outcome of interest was whether delayed transfer to the ICU results in increased morbidity or mortality and the resulting implications on patient health. Other relevant outcomes include measures of disease severity, quality of life, or economic factors like cost-effectiveness. Furthermore, when examining the role of blood gas components in critical illness, outcomes included measures of mortality risk. Statistical significance was described as p <0.05, such as whether high serum anion gap, low pH, elevated lactate and base excess, or low bicarbonate levels are independent predictors of increased mortality. Overall, the systematic assessment of EWS tools, blood gas components were a key focus area for researchers in this field.

## Results

Database searches resulted in 5,865 articles (Figure 1). After removing duplicates followed by title and abstract screening, 454 manuscripts met full-text review criteria. The qualitative review of the studies identified was done by applying the methods are outlined in Table 1. Table 1 further lists the key study characteristics and findings of the study. The review of these studies revealed that delays are due to a combination of lack of ICU resources, flaws in communications among healthcare professionals, and inconsistent transfer guidelines, among others. These transfer delays were associated with poorer clinical outcomes in this high-risk subpopulation. Delays in ICU transfers also led to worse economic outcomes for healthcare institutions. The authors acknowledge that the lack of blinding in several of the analyzed studies could introduce measurement bias in the interpretations.

**Table 1:** Study characteristics, expected study outcomes, and study results.

| Author (year) | Study Design/Country | Study Population | Expected Study Outcome | Results |
| --- | --- | --- | --- | --- |
| Chalfin et al. (2007)<sup>9</sup> | Cross-sectional Analytical Study / US | 50,322 critically ill patients | Delayed transfer (>6 hours) results in increased mortality | Delayed transfer patients (>6hrs) experienced a longer median length of stay (7.0 vs. 6.0 days), an increased ICU mortality rate (17.4% vs 12.9%), and an increased in-hospital mortality rate (17.4% vs 12.9%) |
| Churpek et al. (2017)<sup>1</sup> | Observational Cohort Study / US | 3,789 patients with eCART $\geq 60$ threshold for critical illness | Delayed ICU transfer has a direct correlation with increased mortality rate | Delayed transfer >6 hours was associated with increased mortality compared with early transfer (33.2% vs. 24.5%, $p < 0.001$) |
| Dünser et al. (2024)<sup>4</sup> | Observational Cohort Study / Austria | 34 studies on critically ill ward patients | Delay in ICU admission is associated with increased morbidity and mortality | 4-fold increase in hospital mortality for delayed transfer |
| Franklin et al. (1994)<sup>10</sup> | Retrospective Study / US | 21,505 cardiac arrest patients | Cardiac arrest mortality rate is lowered by rapid ICU transfer | In 99/150 cases, the cardiac arrest was predicted within 6 hours of event |
| Gallo et al. (2024)<sup>2</sup> | Cohort Study / US | 9,938 patients hospitalized in one academic hospital | Implementation of AI deterioration model reduces the risk of escalations among hospitalized patients | 963 patients who met the AI deterioration model's criteria for critical illness experienced a 10.4% risk reduction for their primary outcome at the criteria threshold ($p = 0.03$) |
| Hovmand et al. (2023)<sup>11</sup> | Retrospective Observational Study / Denmark | 369 patients with CABM<sup>a</sup> | Treatment delay associated with increased 30-day mortality | 61% CABM patients had treatment delay >2 hours |
| Huang et al. (2024)<sup>12</sup> | Retrospective Cohort Study / US | 2,774 acute heart failure patients | High serum anion gap indicates increased risk of mortality | Patients with serum anion gap level $\geq 16$ mmol/L have significantly shorter survival times |

<sup>a</sup> CABM, Community Acquired Bacterial Meningitis.

|  |  |  |  |  |
| --- | --- | --- | --- | --- |
| Kiekkas et al. (2022)<sup>14</sup> | Systematic Review / US | 356,936 critically ill adults | Delayed ICU admission is associated with mortality of adult patients | Mortality rate was significantly higher in delayed ICU admission groups in 22 studies |
| Levin et al. (2001)<sup>15</sup> | Cohort Study / US & Canada | 1,332 inpatients with Community-acquired Pneumonia | Minimal use of ABG to manage critically ill patients | Range of ABG use for inpatient was 49.2% to 77.3% |
| Ross et al. (2017)<sup>16</sup> | Retrospective Cohort Study / US | 593 patients at Level I Trauma Center | pH is independent predictor of mortality | Patients with pH <7.0 are 6x less likely to survive |
| Sayers et al. (2016)<sup>17</sup> | Retrospective Observational Cohort Study / US | 3,789 patients in 5 different med-surg units | Delayed ICU transfer increases in-hospital mortality | Patients with delayed transfer had increased length of stay and in-hospital mortality (33.2% vs. 24.5%, p<0.001) |
| Schork et al. (2021)<sup>18</sup> | Retrospective Analysis / Germany | 4,067 ICU surgical patients | Lactate and base excess are strong predictors of mortality | 22% mortality rate in patients with high lactate levels and low base excess levels |
| Talbott et al. (2024)<sup>19</sup> | Retrospective Cohort Study / US | 92 million patients from 55 healthcare organizations | Lower bicarbonate levels are associated with increased mortality | Bicarbonate level ≤5 mEq/L had 6.806 relative risk of death; 6-10 mEq/L had 8.651 risk of death |
| Villar et al. (2019)<sup>20</sup> | Retrospective Cohort Study / US | 3,325 inpatients at US Veteran's Affair Hospital | Serum lactate levels predict risk of death | Patients with median lactate level ~1.7 mmol/L have 2.5% increase in 3-day mortality rate |
| Wrotek et al. (2021)<sup>21</sup> | Retrospective Cohort Study / Poland | 485 children diagnosed with bronchiolitis who had CBG<sup>b</sup> performed | pCO<sub>2</sub> levels were significantly higher in higher risk ICU patients | pCO<sub>2</sub> significantly higher in ICU transferred patients (median 44.8 mmHg vs 36.2 mm Hg, p<0.001) |
| Young et al. (2003)<sup>22</sup> | Inception Cohort Study / US | 91 inpatients with noncardiac diagnoses | Delayed transfer (>4 hours) to ICU is associated with increased mortality | Slow-transfer patients (>4hrs) have increased mortality in-hospital (41% vs 11%) |

<sup>b</sup> CBG, Capillary Blood Gas.

### Impact of ICU transfer delays on clinical outcomes and patient recovery

A comprehensive review of 454 studies has shown that delays in transferring patients from inpatient wards to the ICU are consistently associated with increased in-hospital mortality and an overall negative impact on patient recovery. These delays result in missed opportunities for early intervention and contribute to the deterioration of critically ill patients. In a retrospective observational cohort study, Churpek et al. (2017) analyzed 3,789 patients with eCART scores ≥60—a threshold indicating critical illness—and found a significant increase in in-hospital mortality among patients whose transfer to the ICU was delayed beyond six hours after crossing the threshold. Specifically, patients transferred without delay had a 24.5% in-hospital mortality rate, compared to a 33.2% rate for those who experienced a delay greater than six hours (p < 0.001). While delays persisted despite the use of scoring tools, the implementation of a metric such as eCART improved the overall rate of transfers, suggesting that data-driven risk identification facilitates faster and more efficient clinical decision-making.

Supporting this, a cohort study conducted at a community hospital in Ogden, Utah, found a similar pattern. Researchers defined “slow-transfers” as patients who were transferred to the ICU more than four hours after clinical deterioration. In-hospital mortality among slow-transfers was 41%, compared to just 11% for patients transferred within four hours (Young et al., 2003). This stark contrast demonstrates that even short delays under six hours can significantly increase the risk of death. As summarized in a systematic review by Kiekkas et al. (2022), such delays have been “found to be associated with significantly higher mortality of adult patients,” with 22 studies confirming this trend in a sample of over 356,000 critically ill adults. Similarly, a study by Chalfin et al. (2007) involving 50,322 critically ill patients in the U.S. showed that delays greater than six hours led to an increased ICU mortality rate (17.4% vs. 12.9%), and an increased in-hospital mortality rate (17.4% vs. 12.9%). These results underscore the severe consequences of delayed ICU access—not just on patient survival, but also on resource utilization and system efficiency. A more recent observational cohort study by Dünser et al. (2024), which reviewed 34 studies on critically ill ward patients, revealed a fourfold increase in hospital mortality associated with delayed ICU admission. These findings affirm that early ICU transfer is critical to patient outcomes and should be prioritized in all clinical settings. While not every delay results in death, many have a measurable negative effect on patient recovery and outcomes. The same Ogden study also found that “50% of cardiopulmonary arrests on general medical and surgical wards could be prevented by earlier transfer to ICU” (Young et al., 2003). Timing is a crucial factor in the recovery from acute events such as myocardial infarction, stroke, or respiratory failure. Delayed interventions can mean the difference between full recovery and long-term morbidity.

In a 1994 study spanning a 20-month period, researchers examined 150 in-hospital cardiac arrest events in a public hospital. Of these, 99 patients had shown documented clinical deterioration within six hours of arrest but had not been transferred to the ICU in time (Franklin et al., 1994). The primary causes cited for the delay were poor communication among hospital staff and the absence of a standardized clinical metric to guide ICU admissions. Collectively, the evidence from multiple studies highlights a clear and consistent message: delays in ICU transfer from inpatient units lead to significantly higher mortality, prolonged recovery, and unnecessary strain on healthcare resources.

### Effect of ICU transfer delays on economic outcomes

In addition to numerous adverse clinical impacts, ICU transfer delays impose a significant economic burden on hospitals. A review of 184 studies highlighted the numerous negative outcomes associated with transfer delays, while a broader review of 454 studies demonstrated that such delays from inpatient units to the ICU are linked to increased patient length of stay (LOS) and higher in-hospital mortality rates.

An analytical study using the Project IMPACT database—comprising U.S. ICU patient data from 2000 to 2003—revealed a direct relationship between transfer delays and increased LOS. According to this study, patients transferred to the ICU within six hours had a median LOS of 6.0 days, whereas those whose transfers were delayed for six hours or more had a median LOS of 7.0 days (Chalfin et al., 2007). With over 50,000 patient admissions analyzed, the findings clearly indicated that critically ill patients who experienced transfer delays remained hospitalized longer than those who were promptly transferred. This prolonged stay resulted in the increased utilization of hospital resources, including staff time, ICU resources—ultimately preventing timely care for other critically ill patients.

Delays in ICU transfer not only extended patient LOS but also contributed to a greater consumption of hospital resources. A recent 2024 study by Dunser et al. found that prolonged boarding times for critically ill patients on general wards led to more severe organ dysfunction, greater resource utilization, and longer ICU stays. The longer a patient waits for ICU admission, the more their condition may deteriorate, necessitating more intensive care and extending their time in the ICU. These delays thus translate directly into higher healthcare costs and reduced operational efficiency.

Supporting this, a retrospective analysis conducted in 2010 showed that transfer delays of more than 12 hours were associated with a 12.4% increase in patient LOS. This increase resulted in more than $2 million in additional hospital costs annually (Huang et al., 2010). These findings underscore the importance of improving patient flow and minimizing ICU transfer delays—not only to enhance clinical outcomes but also to reduce unnecessary healthcare expenditures and allow more effective allocation of scarce hospital resources.

### Role of EWS

Several EWS tools are currently in use and have undergone testing (Table 3) with variable results. Among them, NEWS has been implemented nationally by the NHS with some success (Kostakis et al., 2021; Williams, 2022). Furthermore, the effectiveness of EWS tools can be significantly enhanced by incorporating additional factors (Carr et al., 2021). For instance, risk stratification improved significantly when readily available blood and physiological parameters measured at hospital admission were included. A supplemented model incorporating eight routinely collected variables—such as oxygen flow rate, urea, age, C-reactive protein, glomerular filtration rate, neutrophil count, and the neutrophil-to-lymphocyte ratio—showed improved predictive performance (Gallo et al., 2024). A comparative study of six commonly used EWS tools found that NEWS was the most accurate in predicting the risk of death or ICU admission within 24 hours of arrival at the emergency department (Covino et al., 2023).

**Table 2:**
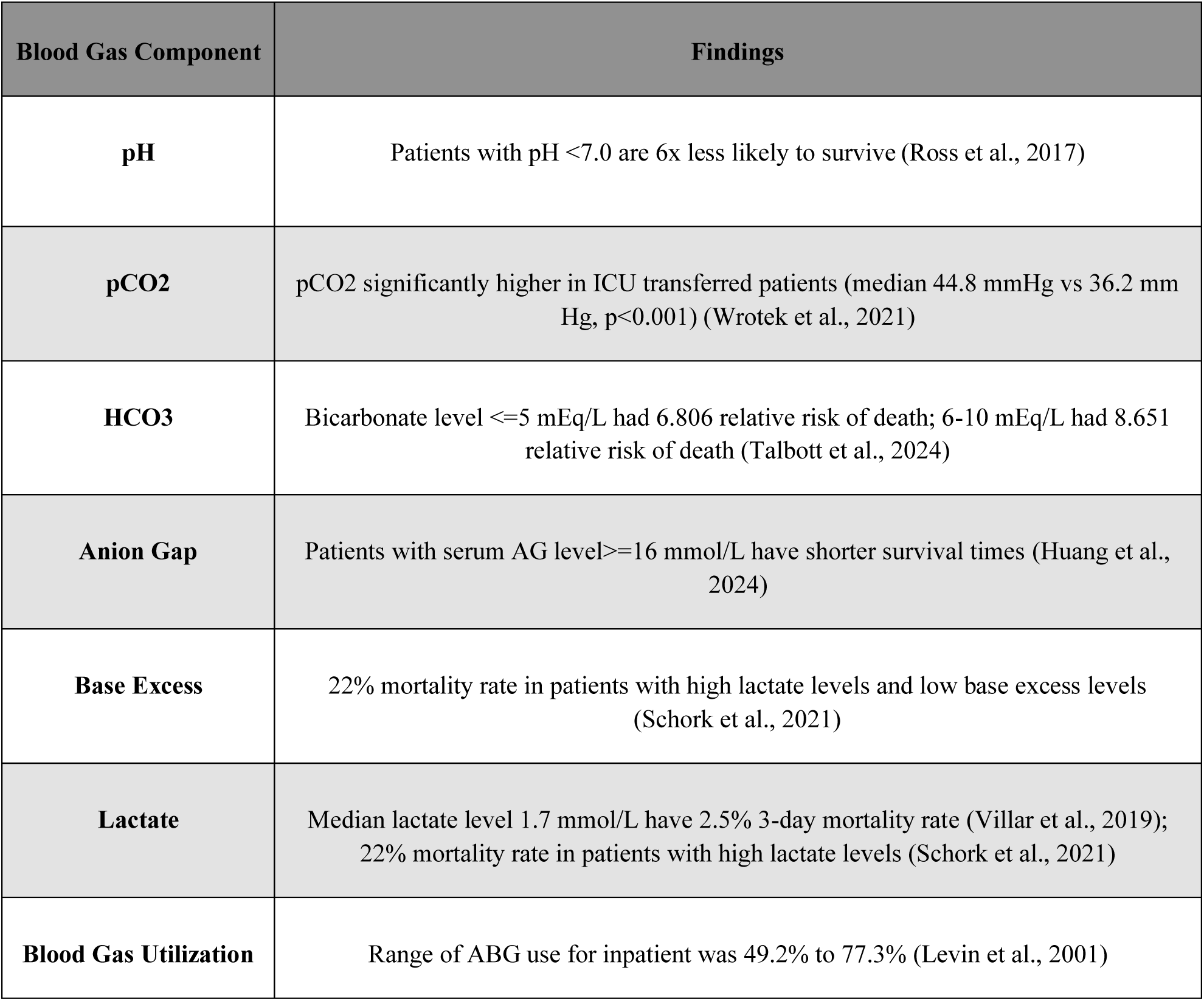
Summary table of blood gas components as independent clinical predictors.

| Blood Gas Component | Findings |
| --- | --- |
| **pH** | Patients with pH <7.0 are 6x less likely to survive (Ross et al., 2017) |
| **pCO<sub>2</sub>** | pCO<sub>2</sub> significantly higher in ICU transferred patients (median 44.8 mmHg vs 36.2 mm Hg, p<0.001) (Wrotek et al., 2021) |
| **HCO<sub>3</sub>** | Bicarbonate level ≤5 mEq/L had 6.806 relative risk of death; 6-10 mEq/L had 8.651 relative risk of death (Talbot et al., 2024) |
| **Anion Gap** | Patients with serum AG level ≥16 mmol/L have shorter survival times (Huang et al., 2024) |
| **Base Excess** | 22% mortality rate in patients with high lactate levels and low base excess levels (Schork et al., 2021) |
| **Lactate** | Median lactate level 1.7 mmol/L have 2.5% 3-day mortality rate (Villar et al., 2019); 22% mortality rate in patients with high lactate levels (Schork et al., 2021) |
| **Blood Gas Utilization** | Range of ABG use for inpatient was 49.2% to 77.3% (Levin et al., 2001) |

**Table 3:** Summary of EWS studies and their pertinent findings.

| Author, Year | EWS related Summary |
| --- | --- |
| **Oneil SM et al. (2021)<sup>3</sup>** | EWS success depends on barriers and facilitators that can be grouped into five overarching themes: Governance, RRT Response, Professional Boundaries, Clinical Experience, and EWS parameters.<br>Barriers: Lack of Standardization, Resources, Lack of accountability, RRT behaviors, Fear, Hierarchy, Increased Conflict, Over confidence, Lack of confidence, and Patient variability.<br>Facilitators: Accountability, Standardization, Resources, RRT behaviors, Expertise, Additional support, License to escalate, Bridge across boundaries, Clinical confidence, empowerment, Clinical judgment, and a tool for detecting deterioration. These are all individual yet inter-related barriers and facilitators to escalation. |
| **Covino M et al. (2023)<sup>5</sup>** | NEWS demonstrates superior accuracy in predicting death or ICU admission within 24 hours of emergency department arrival. It effectively assesses patient risk by comprehensively evaluating key physiological parameters, including respiration rate, oxygen saturation, supplemental oxygen needs, temperature, systolic blood pressure, heart rate, and level of consciousness using the AVPU<sup>c</sup> scale, with reliable calibration across risk levels. |
| **Williams B (2022)<sup>6</sup>** | Ten years' experience of NHS using NEWS/NEWS2, after pioneering a groundbreaking global standard for assessing clinical illness severity through NEWS/NEWS2. By establishing a universal "common language" of early warning indicators—including respiratory rate, oxygen saturation, blood pressure, heart rate, consciousness, temperature, and supplemental oxygen use—the system has become an internationally recognized model for detecting and managing acute patient deterioration. |

<sup>c</sup> AVPU, Alert, Verbal, Painful, Unresponsive

|  |  |
| --- | --- |
| **Gielen A et al. (2021)<sup>7</sup>** | High-risk Early Warning Scores correlate with increased in-hospital mortality, ICU admission, and longer hospital stays, emphasizing the critical importance of precise EWS documentation and monitoring to predict and potentially improve patient outcomes. |
| **Guan G et al. (2022)<sup>23</sup>** | EWS scores' predictability of clinical deterioration is enhanced when the score is applied to patients treated in the hospital setting. However, they are not very effective in prehospital setting. |
| **Muralitharan S et al. (2021)<sup>24</sup>** | ML-based early warning systems can achieve greater accuracy than aggregate-weighted early warning systems. These ML models have the potential to provide clinical decision support however pending rigorous evaluation. |
| **Baker KF et al. (2021)<sup>25</sup>** | NEWS2 monitoring is a sensitive method to identify deterioration of hospitalized COVID-19 patients. The downside is high false-trigger rate. |
| **Eddahchouri Y et al. (2022)<sup>26</sup>** | Wearable monitoring technology providing continuous monitoring of patient vital signs wirelessly to hospital systems was associated with a reduction in unplanned ICU admissions and rapid response calls. |
| **Kostakis I et al. (2021)<sup>27</sup>** | NEWS2 is a standardized scoring system that utilizes six routinely collected physiological parameters to assess acute illness severity and risk of deterioration: respiration rate, oxygen saturation, systolic blood pressure, pulse rate (heart rate), level of consciousness or new confusion, and body temperature. Each parameter is assigned a score based on how much it deviates from the normal range, and these individual scores are aggregated to generate a total NEWS2 score ranging from 0 to 20. Additionally, patients requiring supplemental oxygen receive an extra 2 points added to their score.<br>Support for the use of NEWS or NEWS2 as national and international recommendations for the assessment of acute-illness severity in patients with COVID-19. |
| **Mcgaughey J et al. (2021)<sup>28</sup>** | Low to moderate evidence that EWS and RRS may lead to little or no difference in hospital mortality, unplanned ICU admissions, length of hospital stay or adverse events, and little to no difference in composite outcomes. This highlights the diversity in outcomes due to varying quality of most studies investigating EWS and RRS. |
| **Romero-Brufau S et al. (2021)<sup>29</sup>** | MC-EWS<sup>d</sup> demonstrated advanced predictive capabilities for inpatient deterioration by integrating physiological parameters to improve clinical risk assessment and reduce unnecessary alerts. Specifically, by integrating parameters such as age, oxygen saturation to inspired oxygen ratio, respiratory rate, white cell count, venous lactate, heart rate to systolic blood pressure ratio, albumin, oxygen saturation, urea, and heart rate. Secondary variables derived from shock index, Braden score, respiratory rate, and oxygen saturation and respiratory index with oxygen, along with systolic blood pressure and heart rate. |
| **Carr E et al. (2021)<sup>30</sup>** | NEWS2 was enhanced for prediction of severe COVID-19 outcomes by including blood and physiological parameters such as supplemental oxygen flow rate, urea, age, oxygen saturation, C-reactive protein, estimated glomerular filtration rate, neutrophil count, neutrophil/lymphocyte ratio. |
| **Adams R et al. (2022)<sup>31</sup>** | In this prospective, multi-site cohort study of 590,736 patients, sepsis EWS called the TREWS identified sepsis patients early and improved patient outcomes. |

<sup>d</sup> MC-EWS, Machine learning-enhanced Early Warning Score.

A cohort study by Gallo et al. (2024) also found that the implementation of an artificial intelligence (AI)-enabled deterioration model was associated with a significant reduction in care escalations among inpatients. This model utilized vital signs, laboratory results, and demographic data to calculate a real-time risk score for cardiac arrest. Machine learning-based EWS models have been shown to outperform traditional aggregate-weighted systems and could be considered the frontier. For example, Muralitharan et al. (2021) demonstrated that machine learning approaches yielded greater predictive accuracy. Similarly, Romero-Brufau et al.’s (2021) model which incorporated variables such as the oxygen saturation-to-inspired oxygen ratio, respiratory rate, white cell count, venous lactate, heart rate-to-systolic blood pressure ratio, albumin, oxygen saturation, urea, and heart rate enhanced predictive capabilities.

In a large, prospective, multi-site cohort study, the Targeted Real-time Early Warning System (TREWS), a sepsis alert system, monitored 590,736 patients and successfully identified sepsis cases early, leading to improved outcomes. It was particularly effective in identifying patients most likely to benefit from early intervention (Adams et al., 2022).

Despite these advancements, the success of EWS tools can vary depending on the clinical setting, calibration, and human error (Gielen et al., 2021). McGaughey et al. (2021) found that EWS and rapid response systems (RRS) may lead to little or no significant differences in hospital mortality, unplanned ICU admissions, length of stay, or adverse events, highlighting the variability in outcome selection and methodological quality across studies. Moreover, Guan et al. (2022) observed that while EWS performs better for patients already in the hospital, its high thresholds and poor predictive value for 30-day mortality make it less effective in prehospital settings. Additionally, the system can have a relatively high false-trigger rate (Baker et al., 2021) leading to excessive alerts and ‘alarm fatigue’.

Emerging evidence suggests that for tracking vital signs, prehospital blood gases might help reduce mortality, though the evidence remains limited. For example, continuous vital sign monitoring through wearable technology, wirelessly linked to hospital systems, has been associated with reductions in unplanned ICU admissions and RRT calls (Eddahchouri et al., 2022) albeit at the expense of a high false-trigger rate.

### BGA as a biomarker

BGA is a common test that quantifies gas pressures and other relevant blood characteristics that are time-stamped to assess a patient’s clinical status. Arterial blood gas (ABG) and venous blood gas (VBG) measurements can be used interchangeably in many clinical contexts, apart from specific parameters such as partial pressure of oxygen (pO₂), for which arterial sampling remains essential. VBG sampling is easier to perform, less painful for patients, and associated with fewer procedural risks, and these advantages have contributed to a growing preference for VBG in routine clinical practice. Commonly measured parameters in BGA include pH, partial pressures of oxygen (pO₂) and carbon dioxide (pCO₂), bicarbonate (HCO₃), lactate, serum anion gap (AG), and base excess, among others. These could be relevant as noted in Romero-Brufau et al.’s (2021) EWS model which incorporated variables such as the oxygen saturation-to-inspired oxygen ratio, respiratory rate, venous lactate, oxygen saturation.

In a retrospective cohort study conducted at a Level I trauma center in the U.S., pH was identified as an independent predictor of inpatient mortality. The study found that patients with a pH lower than 7.0 were six times less likely to survive, and those with a pH lower than 6.7 were twelve times less likely to survive (Ross et al., 2017). This indicates that trauma patients, even in a severely acidotic state, may benefit from rapid ICU transfer to expedite treatment. Similarly, a retrospective study of 485 children (ages 8 days to 22 months) diagnosed with bronchiolitis found that elevated pCO₂ levels were significantly associated with ICU admission. Patients transferred to the ICU had a median pCO₂ of 44.8 mmHg compared to 36.2 mmHg in those who were not, with the difference being statistically significant (p < 0.001) (Wrotek et al., 2021). These findings suggest that pCO₂ may be a valuable marker for assessing ICU risk in pediatric respiratory conditions. HCO₃ levels have also emerged as a strong prognostic indicator. Talbott et al. (2024), in a large-scale retrospective cohort study involving over 92 million trauma patients from 55 healthcare organizations, found that lower HCO₃ levels were significantly associated with increased mortality. Patients with HCO₃ ≤5 mEq/L had a relative risk of death of 6.806, while those with levels between 6–10 mEq/L had a risk of 8.651, underscoring its predictive power in trauma care. Lactate, another key measurement in BGA, typically increases with tissue hypoxia and is widely recognized as a sepsis predictor (Villar et al., 2019). In a retrospective analysis of 4,067 ICU surgical patients in Germany, researchers found that lactate level was the strongest predictor of mortality. The study reported a 22% mortality rate among patients with elevated lactate and reduced base excess levels. It also found that the minimum base excess level had a strong correlation with mortality, particularly in post-cardiac surgery patients (Schork et al., 2021). The AG is another important BGA parameter. Huang et al. (2024) conducted a retrospective cohort study involving 2,774 patients with acute heart failure (AHF) and concluded that high serum AG at admission was associated with worse outcomes. Patients with AG ≥16 mmol/L had significantly shorter survival times and poorer recovery, indicating that elevated AG is a marker of rapid clinical deterioration. Collectively, BGA components—pH, pCO₂, HCO₃, lactate, base excess, and AG—demonstrate predictive value in determining a patient’s prognosis, time to intervention and resource needs. Additionally, given BGA’s already widespread use in clinical practice (Levin et al., 2001), its expanded application as an adjunct to supplement EWS tools would be clinically acceptable.

## Discussion

ICU transfer delays are a pervasive problem impacting patient outcomes for patients requiring transfer to ICU during their hospital stays. A study by Churpek et al. (2016) found that for each additional hour of delay in ICU admission, there was a 3% increase in the odds of mortality. This underscores the urgent need for rapid escalation of care—every hour of delay contributes incrementally and significantly to worse patient outcomes. Studies included in this systematic review define a “significant transfer delay” as a duration equal to or greater than six hours. While this six-hour benchmark is widely cited, it should not be interpreted as a strict threshold. Currently, many hospitals rely on subjective assessments to determine ICU transfer needs, based on a combination of clinical status, laboratory results, and vital signs. These decisions are often further influenced by process and human factors such as the judgment of the accepting physician.

Review of literature supports the use of EWS tools with intra-hospital transfer. There is moderate evidence supporting use of tools like the EWS, eCART, and Machine Learning (ML) algorithms which have improved risk stratification. However, they are not without limitations. eCART, for instance, utilizes a combination of physiological and laboratory parameters—including heart rate, respiratory rate, blood pressure, oxygen saturation, temperature, electrolytes, blood urea nitrogen, creatinine, white blood cell count, hemoglobin, age, and gender—to predict adverse outcomes like cardiac arrest. In summary, their effectiveness is contingent on the presence of a robust electronic medical record (EMR) infrastructure and susceptibility to human factors and needs to be calibrated for the institution.

Furthermore, EWS tools can be enhanced by using an objective, time-stamped metric—such as BGA results to augment decision-making in real time. BGA provides rapid insights into physiological stability by measuring pH, pCO₂, pO₂, HCO₃⁻, lactate, and other parameters closely tied to patient deterioration. Its predictive capability has been demonstrated in numerous studies and could allow EMRs to automatically flag at-risk patients for early ICU evaluation.

## Conclusion

As hospitals develop strategies to address delays in escalation of care, they must consider several interconnected factors, including accountability, standardization, resource availability, the behavior and responsiveness of RRT teams, clinical expertise, confidence and judgment, and empowerment of frontline staff. O’Neill et al. (2021) emphasized that these elements are often institution specific and may function as both barriers and facilitators to effective escalation, and they must hence be addressed in parallel with the implementation of EWS tools.

Overall, the cumulative evidence clearly demonstrates that ICU transfer delays are harmful, leading to increase in-hospital mortality and unnecessary strain on healthcare resources. Even relatively brief delays of four to six hours can have severe clinical consequences. Therefore, timely recognition of clinical deterioration, consistent communication, and the use of objective, predictive tools such as EWS, NEWS, and eCART are vital. These tools need to be calibrated to the institutions to align with local workflows. For instance, high rate of false triggers would require thresholds to be calibrated locally. Furthermore, EWS can be enhanced by incorporating certain metrics such as BGA. The addition of BGA as a supplemental metric could enhance the precision of these tools and improve care for the most vulnerable patient populations.

Hospitals must prioritize the early identification of patients who require critical care, minimize transfer delays, and establish standardized, team-based communication protocols. Embedding EWS tools, enhancing them with BGAs, calibrating to the institution, using real-time EMR alerts, and automated monitoring into clinical workflows will be essential for preventing avoidable deterioration. Ultimately, optimizing ICU transfer processes is not merely a clinical goal—it is an ethical obligation and an operational necessity in delivering timely, high-quality care.

## Data Availability

All data produced in the present work are contained in the manuscript.

## Author Contributions

**Praveen Meka**: Conceptualization, Software, Validation, Resources, Writing – Review & Editing, Supervision, Project administration**. Emma Stiller**: Formal analysis, Investigation, Resources, Data Curation, Writing – Original Draft, Writing – Review & Editing.

The authors report no proprietary or commercial interest in any product mentioned or concept discussed in this article. No funding was received for this study.

